# Evaluation of SARS-CoV-2 in Breastmilk from 18 Infected Women

**DOI:** 10.1101/2020.06.12.20127944

**Authors:** Christina D. Chambers, Paul Krogstad, Kerri Bertrand, Deisy Contreras, Nicole H. Tobin, Lars Bode, Grace M. Aldrovandi

**Affiliations:** University of California, San Diego, 9500 Gilman Drive MC0828, La Jolla, CA 92093-0828; University of California, Los Angeles, 10833 Le Conte Ave, 22-442 MDCC, Los Angeles, CA 90095; University of California, San Diego, 9500 Gilman Drive MC0715, La Jolla, CA 92093-0715

## Abstract

Currently, the U.S. Centers for Disease Control and Prevention, American Academy of Pediatrics and the World Health Organization advise that women who are infected with SARS-CoV-2 may choose to breastfeed with appropriate protections to prevent transmission of the virus through respiratory droplets.^(1)^ However, the potential for exposure to SARS-CoV-2 through breastfeeding is currently unknown. To date, case reports on breastmilk samples from a total of 24 SARS-CoV-2-infected women have been published.^(2-7)^ Of those, viral RNA was detected in ten breastmilk samples from four women. In some but not all cases, environmental contamination as the source of the virus or retrograde flow from an infected infant could not be ruled out.

---

We established a quantitative RT-PCR assay for SARS-CoV-2 in breastmilk with a limit of detection of 250 copies per mL and validated it by spiking breastmilk from uninfected women with known amounts of viral RNA. In addition, we established tissue culture methods to detect replication-competent SARS-CoV-2 in breastmilk. No viral RNA nor culturable virus was detected after Holder pasteurization of breastmilk samples that had been spiked with replication-competent SARS-CoV-2 (see Supplement).

Between March 27 and May 6, 2020, we collected and analyzed 64 serial breastmilk samples from 18 SARS-CoV-2-infected women residing in the U.S. (see Supplement for clinical characteristics). Breastmilk samples were collected before and after women had a positive SARS-CoV-2 RT-PCR test and all but one woman had symptomatic disease (see Figure). One of the 64 breastmilk samples had detectable SARS-CoV-2 RNA by RT-PCR. The positive sample was collected on the day of symptom onset but one sample 2 days prior to symptom onset and two subsequent samples, collected 12 and 41 days later, tested negative for viral RNA. In addition, a subset of 26 breastmilk samples from nine women were tested for the presence of replication-competent virus using our established culture methods, and all were negative including the one sample that tested positive for viral RNA by RT-PCR.

**Figure.**
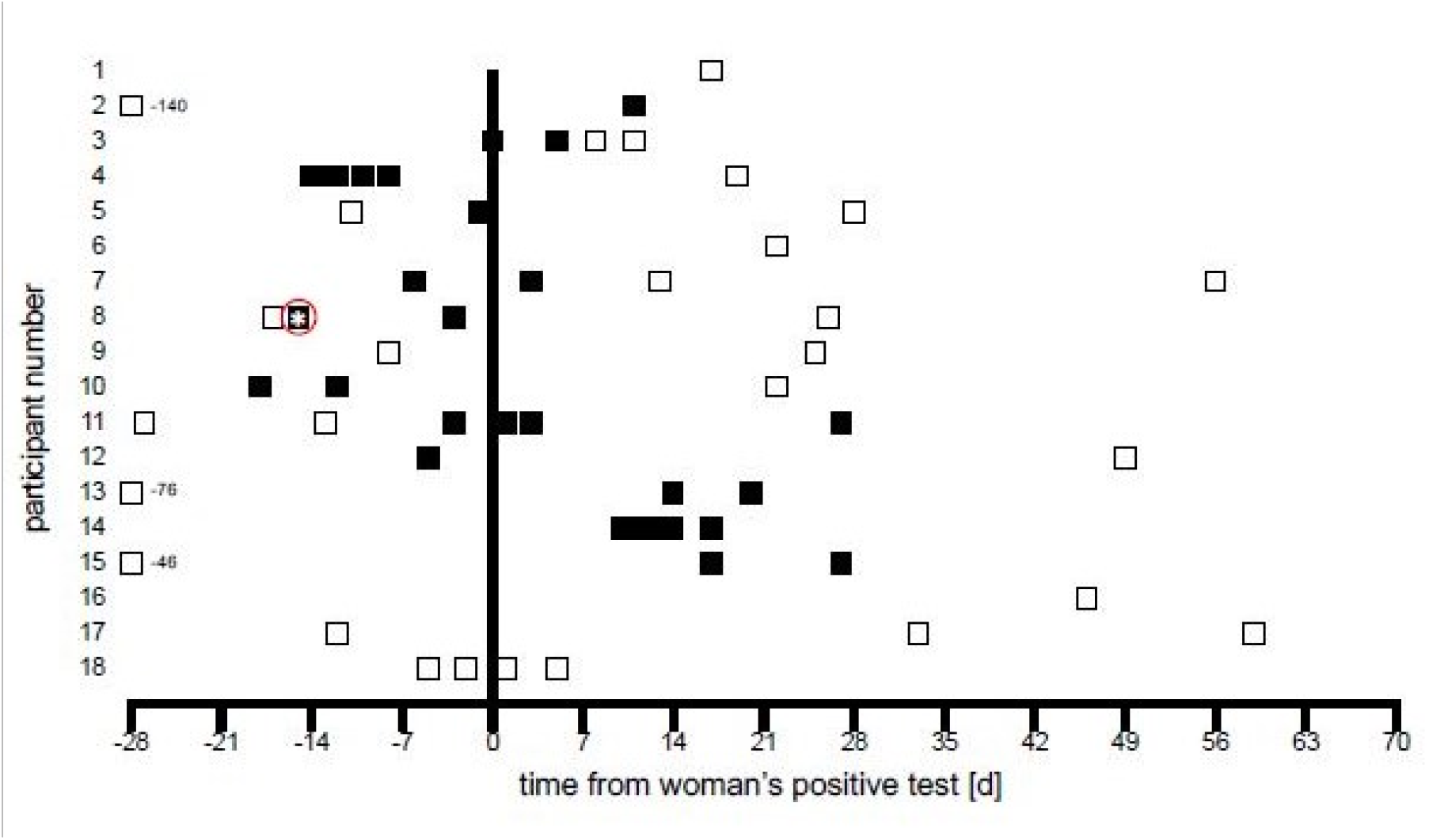
Breastmilk Sampling Relative to Time of Woman’s Positive SARS-CoV-2 Test. Filled and unfilled boxes indicate breastmilk samples that were collected when the woman was symptomatic and asymptomatic, respectively. All samples were tested for SARS-CoV-2 viral RNA by PT-PCR. Samples from participants 1-10, excluding participant 3, were also tested in infectivity assay. The sample highlighted by asterisk tested positive by RT-PCR, but negative by infectivity assay.

Although SARS-CoV-2 RNA was detected in one milk sample from one of eighteen infected women, the viral culture for that sample was negative. This suggests that SARS-CoV-2 RNA does not represent replication-competent virus and that breastmilk itself is likely not a source of infection for the infant. Furthermore, when control breastmilk samples spiked with replication-competent SARS-CoV-2 virus were treated by Holder pasteurization, a process commonly performed by donor milk banks, no replication-competent virus nor viral RNA was detectable. Further research to confirm these findings is needed, as well as an examination of convalescent milk for the presence of antibodies against SARS-CoV-2.

## Data Availability

Data access may be provided, with appropriate ethics approval, by contacting the authors.

## Evaluation of SARS-CoV-2 in Breastmilk from 18 Infected Women Supplementary Appendix

### Methods

#### Clinical data and sample collection

Women residing in the U.S. or Canada were invited to enroll in the Mommy’s Milk Human Milk Research Biorepository at the University of California San Diego. Women were interviewed by telephone about COVID-19 symptoms and SARS-CoV-2 test results. Kits for self-collection of milk were mailed to participants. Breastmilk was collected using their personal breast pump and following standardized instructions for expressing and storing milk samples in provided sterile milk collection bags. Instructions included hand washing before and after milk expression and cleansing of the nipple and areola with an alcohol wipe provided in the kit. Women who had recovered from their illness at the time of the study interview were asked to ship any frozen samples from the peak of their symptoms in addition to a fresh milk sample. All fresh samples were shipped to the Biorepository overnight on ice within twenty-four hours of collection. Samples were then aliquoted and stored at −80°C prior to shipment on dry ice to the University of California Los Angeles. All women provided written informed consent. Institutional review board approval for the study was obtained at the University of California San Diego.

Characteristics of the women and their children, timing of breastmilk sample collection relative to the positive SARS-CoV-2 test and the presence of specific symptoms of the infection are shown in Tables S1 and S2.

#### Detection of SARS-CoV-2 RNA in breastmilk

We established and validated a quantitative RT-PCR assay using the U.S. Food and Drug Administration Emergency Use Authorization approved Abbott m2000sp/rt platform and the Abbott RealTime SARS-CoV-2 Assay. We spiked known amounts of viral RNA into breastmilk samples collected from 30 healthy, uninfected women participating in our studies prior to 2017. We established a limit of detection of 250 copies of SARS-CoV-2 RNA per ml of breastmilk. This assay was then used to test 64 breastmilk samples from 18 women for SARS-CoV-2 RNA.

#### Detection of replication-competent SARS-CoV-2 in breastmilk

In addition to detection of viral RNA, we established tissue culture methods to detect replication-competent SARS-CoV-2 in breastmilk and to determine if factors present in human milk interfere with its infectivity. A stock of SARS-CoV-2 (USA-WA1/2020, BEI Resources, Manassus, VA) was propagated in Vero-E6 green monkey kidney cells and quantified by limiting dilution with viral titers expressed as TCID_50_ (Tissue Culture Infectious Doses). A 20-fold range of SARS-CoV-2 was added to two samples of breastmilk donated by different women before 2017. After four days in Vero-E6 culture, SARS-CoV-2 was detected even at the lowest amount of virus added (100 TCID_50_). This was confirmed in additional samples from eleven different women spiked with 200 TCID_50_. In each case, replication was demonstrated by cytopathic effects in culture and SARS-CoV-2 replication was confirmed by RT-PCR.

#### Culture of breastmilk from women with SARS-CoV-2 infection

Twenty-six breastmilk samples from nine women were cultured in Vero-E6 cells using the methods described above. In all twenty-six samples, cytopathic effects were not observed and RT-PCR of culture supernatants were negative.

#### Effect of Holder Pasteurization

SARS-CoV-2 (200 TCID_50_) was added to breastmilk samples from two different donors, heated to 62.5°C for 30 minutes and then cooled to 4°C to mimic the conditions of Holder pasteurization commonly used in human milk banks. Following this procedure, the samples were added to Vero-E6 cell culture. Viral RNA was not detected by RT-PCR analysis in two of two samples and no cytopathic effect was observed. Non-pasteurized aliquots of the same two milk/virus mixtures were cultured in parallel and viral replication occurred in both samples.

## Results

**Table S1.** Characteristics of Women and Breastmilk Sample Collection for 18 Women Positive for SARS-CoV-2 Infection.

| **Participant Number** | **Age Category (yrs)** | **Race Ethnicity** | **SES Category \*** | **BMI Category \*\*** | **Risk Factors for SARS-CoV-2 Infection** | **Source of Infection** | **Reported SARS-CoV-2 Symptoms** | **Number of Samples Collected N=64** |
| --- | --- | --- | --- | --- | --- | --- | --- | --- |
| 1 | 40+ | Non-Hispanic Caucasian | 1 | 25-29 | None | Unknown | Fever, Body Aches, Headache, Shortness of Breath, Loss of Smell | 1 |
| 2 | 30-34 | Non-Hispanic Caucasian | 1 | 30+ | Asthma, Congenital Kidney Defect | Occupational | Fever, Chills, Body Aches, Headache, Cough, Shortness of Breath, Loss of Smell, Loss of Appetite | 2 |
| 3 | 25-29 | Hispanic Caucasian | 1 | 30+ | Asthma, Polycystic Ovary Syndrome | Household | Fever, Chills, Body Aches, Headache, Sore Throat, Cough, Loss of Smell | 4 |
| 4 | 30-34 | Non-Hispanic Caucasian | 1 | 19-24 | None | Occupational | Fever, Body Aches, Headache, Sore Throat, Diarrhea, Fatigue | 6*** |
| 5 | 35-39 | Non-Hispanic Caucasian | 1 | 19-24 | None | Household | Chills, Body Aches, Headache, Runny Nose, Cough, Diarrhea, Loss of Smell, Loss of Taste, Loss of Appetite | 3 |
| 6 | 25-29 | Non-Hispanic Caucasian | 5 | 19-24 | None | Household | Fever, Body Aches, Headache, Runny Nose, Cough, Shortness of Breath, Diarrhea, Loss of Appetite | 1 |
| **Participant** | **Age** | **Race** | **SES** | **BMI** | **Risk Factors** | **Source of** | **Reported** | **Number** |

| **Number** | **Category**<br>**(yrs)** | **Ethnicity** | **Category**<br>**\*** | **Category**<br>**\*\*** | **for SARS-**<br>**CoV-2**<br>**Infection** | **Infection** | **SARS-CoV-2**<br>**Symptoms** | **of**<br>**Samples**<br>**Collected**<br>**N=64** |
| --- | --- | --- | --- | --- | --- | --- | --- | --- |
| 7 | 35-39 | Non-Hispanic<br>Caucasian | 1 | 25-29 | Endometriosis | Household | Fever, Body<br>Aches,<br>Headache,<br>Runny Nose,<br>Cough,<br>Shortness of<br>Breath,<br>Diarrhea,<br>Loss of<br>Appetite | 4 |
| 8<sup>+</sup> | 25-29 | Non-Hispanic<br>Caucasian | 1 | <19 | Hypertension | Community | Chills, Body<br>Aches, Runny<br>Nose, Sore<br>Throat,<br>Cough,<br>Shortness of<br>Breath, Loss<br>of Smell,<br>Loss of Taste | 4 |
| 9 | 40+ | Non-Hispanic<br>Caucasian | 1 | <19 | None | Occupational | Fever, Chills,<br>Body Aches,<br>Headache,<br>Cough,<br>Nausea/Vomi<br>ting,<br>Diarrhea,<br>Loss of<br>Smell, Loss<br>of Taste, Loss<br>of Appetite | 2 |
| 10 | 35-39 | Non-Hispanic<br>Caucasian | 1 | 19-24 | None | Unknown | Body Aches,<br>Sore Throat,<br>Cough, Loss<br>of Smell | 3 |
| 11 | 30-34 | Non-Hispanic<br>Caucasian | 1 | 25-29 | None | Unknown | Fever, Chills,<br>Body Aches,<br>Headache,<br>Runny Nose,<br>Sore Throat,<br>Cough,<br>Shortness of<br>Breath,<br>Abdominal<br>Pain,<br>Diarrhea,<br>Loss of<br>Smell, Loss<br>of Taste, Loss<br>of Appetite,<br>Insomnia | 6 |
| **Participant**<br>**Number** | **Age**<br>**Category** | **Race**<br>**Ethnicity** | **SES**<br>**Category** | **BMI**<br>**Category** | **Risk Factors**<br>**for SARS-** | **Source of**<br>**Infection** | **Reported**<br>**SARS-CoV-2** | **Number**<br>**of** |

|  | (yrs) |  | * | ** | CoV-2<br>Infection |  | Symptoms | Samples<br>Collected<br>N=64 |
| --- | --- | --- | --- | --- | --- | --- | --- | --- |
| 12 | 40+ | Non-Hispanic<br>Caucasian | 1 | 19-24 | None | Unknown | Fever, Body Aches,<br>Headache,<br>Sore Throat,<br>Cough | 2 |
| 13 | 25-29 | Non-Hispanic<br>Caucasian | 1 | 25-29 | Asthma | Occupational | Fever, Body Aches,<br>Headache,<br>Runny Nose,<br>Sore Throat,<br>Shortness of Breath, Loss<br>of Appetite | 3 |
| 14 | 35-39 | Non-Hispanic<br>Native American | 3 | ≥30 | Type II Diabetes,<br>Hypertension | Occupational | Fever, Chills,<br>Body Aches,<br>Headache,<br>Runny Nose,<br>Cough,<br>Shortness of Breath,<br>Diarrhea,<br>Loss of Smell, Loss<br>of Taste, Loss<br>of Appetite,<br>Hospitalized | 12 |
| 15 | 35-39 | Hispanic<br>Caucasian | 1 | ≥30 | Hypertension,<br>High Cholesterol | Household | Fever, Chills,<br>Body Aches,<br>Headache,<br>Runny Nose,<br>Sore Throat,<br>Nausea/<br>Vomiting,<br>Diarrhea,<br>Loss of Smell, Loss<br>of Taste, Pain<br>in Ribs/Back | 3 |
| 16 | 35-39 | Non-Hispanic<br>Caucasian | 2 | 25-29 | None | Unknown | Fever, Chills,<br>Body Aches,<br>Headache,<br>Cough,<br>Shortness of Breath,<br>Nausea/Vomiting,<br>Abdominal Pain,<br>Diarrhea,<br>Loss of Smell, Loss | 1 |
| Participant<br>Number | Age<br>Category | Race<br>Ethnicity | SES<br>Category | BMI<br>Category | Risk Factors<br>for SARS- | Source of<br>Infection | Reported<br>SARS-CoV-2 | Number<br>of |

|  | (yrs) |  | * | ** | CoV-2 Infection |  | Symptoms | Samples Collected N=64 |
| --- | --- | --- | --- | --- | --- | --- | --- | --- |
| 16 cont. |  |  |  |  |  |  | of Taste, Loss of Appetite |  |
| 17 | 35-39 | Non-Hispanic Caucasian | 1 | ≥30 | Asthma, Polycystic Ovary Syndrome | Occupational | Fever, Chills, Body Aches, Headache, Cough, Nausea/Vomiting, Loss of Smell, Loss of Taste | 3 |
| 18 | 30-34 | Non-Hispanic Asian | 2 | 19-24 | None | Community | Asymptomatic | 4 |

\*SES = socioeconomic status category calculated using Hollingshead Scale 1-5 based on mother's and father's occupations and education; category 1 highest and category 5 lowest<sup>1</sup>
\*\*BMI – body mass index calculated based on current weight in kilograms divided by height in meters<sup>2</sup>
\*\*\*Milk samples were pumped during the peak of symptoms on days 1-6 after onset of symptoms; however, the participant did not date the individual milk collection bags
+ Woman with detectable SARS-CoV-2 RNA by RT-PCR in one breastmilk sample

**Table S2:** Characteristics of Children of the 18 Women Positive for SARS-CoV-2 Infection.

| Participant Number | Child Age (months) | SARS-CoV-2 Testing Status | Days Since Mother's Onset of Symptoms to Child Onset of Symptoms | Days since Mother's + test to child Diagnostic Test | Symptoms | Symptom Duration (days) | Medical Care |
| --- | --- | --- | --- | --- | --- | --- | --- |
| 1 | 8 | Positive | 0 | 3 | Labored Breathing, Fatigue and Fussy | 12 | Emergency Room |
| 2 | 4 | Not Tested | NA | NA | Asymptomatic | NA | NA |
| 3 | 25 | Not Tested | NA | NA | Asymptomatic | NA | NA |
| 4 | 4 | Not Tested | -4 | NA | Cough and Mild Congestion | 5 | None |
| 5 | <1 | Not Tested | 3 | NA | Diarrhea | 3 | None |
| 6 | 19 | Negative | NA | NA | Cough, Runny Nose | 5 | None |
| 7 | 3 | Not Tested | NA | NA | Asymptomatic | NA | NA |
| 8 | 9 | Not Tested | 2 | NA | Fever | 1 | None |
| 9 | 18 | Presumptive Positive | 3 | NA | Fever, Irritability | 2 | None |
| 10 | 5 | Not Tested | 7 | NA | Fever, Lethargic, Wheezing, Nasal Congestion, Dry Cough | 17 | Emergency Room |
| 11 | 5 | Not Tested | 5 | NA | Fever, Not Sleeping, Fussy | 3 | None |
| 12 | 11 | Not Tested | 2 | NA | Fever | 2 | None |
| Participant Number | Child Age (months) | SARS-CoV-2 Testing Status | Days Since Mother's Onset of | Days since Mother's + test to child | Symptoms | Symptom Duration (days) | Medical Care |

|  |  |  | Symptoms to<br>Child Onset<br>of Symptoms | Diagnostic<br>Test |  |  |  |
| --- | --- | --- | --- | --- | --- | --- | --- |
| 13 | 21 | Presumptive<br>Positive | 10 | NA | Headache, Fatigue | 1 | None |
| 14 | <1 | Positive | 4 | 4 | Asymptomatic | NA | None |
| 15 | 2 | Presumptive<br>Positive | -9 | NA | Fever,<br>Nausea/Vomiting,<br>Breathing Problems,<br>Retractions,<br>Congestion | 22 | Nebulizer,<br>Inhaler |
| 16 | 19 | Not Tested | 11 | NA | Fever, Dry Cough,<br>Runny Nose | 1 | None |
| 17 | 8 | Not Tested | 0 | NA | Fever, Fatigue,<br>Body Aches | 3 | None |
| 18 | 4 | Not Tested | NA | NA | Asymptomatic | NA | NA |

## Acknowledgments

The authors wish to thank the women and their infants who participated in this study, and who gave so generously of their time and effort to provide milk samples and clinical information.

Christina Chambers received an award for this study from the University of California Office of the President Emergency COVID-19 Research Program, and also received resources for the Human Milk Research Biorepository from the Altman Clinical Translational Research Institute (ACTRI) at UC San Diego funded by the National Institutes of Health (NIH) National Center for Advancing Translational Sciences (NCATS) under Award Number UL1TR001442. Medela Corporation provided milk sample collection materials for this study. Costs for shipping of milk samples was financially supported by the Mothers’ Milk Bank at Austin, an accredited milk bank and member of the Human Milk Banking Association of North America.

Lars Bode is the UC San Diego Chair of Collaborative Human Milk Research, endowed by the Family Larsson-Rosenquist Foundation, that also provided an unrestricted COVID19 emergency gift fund. Grace Aldrovandi is supported by the IMPAACT Network. Overall support for the International Maternal Pediatric Adolescent AIDS Clinical Trials Group (IMPAACT) was provided by the National Institute of Allergy and Infectious Diseases (NIAID) of the National Institutes of Health (NIH) under Award Numbers UM1AI068632 (IMPAACT LOC), UM1AI068616 (IMPAACT SDMC) and UM1AI106716 (IMPAACT LC), with co-funding from the *Eunice Kennedy Shriver* National Institute of Child Health and Human Development (NICHD) and the National Institute of Mental Health (NIMH). The content is solely the responsibility of the authors and does not necessarily represent the official views of the NIH. Paul Krogstad received supported from the University of California Los Angeles (UCLA) AIDS Institute, UCLA CFAR (AI028697), the James B. Pendleton Charitable Trust, and the McCarthy Family Foundation.

